# Single-label and Multi-label Classification for Disease Recognition with Special Consideration of Comorbidities

**DOI:** 10.64898/2025.12.23.25342901

**Authors:** Sophie Schmiegel, Hannah Marchi, Marvin-Hendrik Röchter, Martin Rudwaleit, Christiane Fuchs

**Affiliations:** Faculty of Business Administration and Economics, Data Science Group, Bielefeld University, Germany; Institute of Computational Biology, Computational Health Center, Helmholtz Munich, Neuherberg, Germany; Hospital Bielefeld Rosenhöhe, Department of Internal Medicine and Rheumatology; Bielefeld University, Medical School and University Medical Center OWL

**Keywords:** class imbalance, classifier chain (CC), comorbidities, decision space, immune-mediated inflammatory disease (IMID), multi-label classification (MLC), single-label classification (SLC)

## Abstract

Certain diseases require rapid treatment to avoid long-term consequences for patients. However, they may be difficult to recognize, especially if the symptoms are ambiguous and compatible with multiple possible diagnoses. Completing all necessary examinations often takes time, thereby prolonging patient suffering. Data-driven approaches, such as single-label classification (SLC) and multi-label classification (MLC), can help accelerate the diagnostic process and improve accuracy. These two approaches differ primarily in the number of classes they allow a sample to belong to: SLC assumes the classes being mutually exclusive so that each sample belongs to exactly one class whereas MLC supposes the classes being mutually inclusive, i.e. a sample can belong to several classes or none, acknowledging the possibility of comorbidities. Comparing SLC and MLC allows us to investigate whether disease recognition benefits from considering comorbidities. In this context, we aim to provide a conceptual framing of (differences between) the two approaches in model formulation, decision spaces and handling of class imbalance. To empirically assess their performance, we conduct a case study applying SLC and MLC to data from chronic pain patients. Our analysis yields an ambiguous picture of whether incorporating comorbidities improve disease recognition. The suitability of SLC and MLC is determined by multiple factors, notably the dependency structure among diseases and between diseases and covariates, as well as by data characteristics such as class imbalance. This highlights the importance of considering the specific characteristics of the data when selecting an appropriate classification approach for disease recognition and beyond.

## 1 Introduction

Patients can suffer from several diseases simultaneously. With regard to relevant symptoms, there is often one disease that is mainly responsible. This disease is called ‘main disease’ in the following. However, possible other diseases, known as ‘comorbidities’, might be present and cause similar symptoms. Based on this knowledge, various differential diagnoses are conceivable. Investigations to verify all suspected diagnoses may take several weeks or months, especially if waiting times for appointments at secondary care units are long. This can lead to a delay in adequate treatment prolonging the patient’s suffering and potentially leading to long-term consequences. It seems reasonable to consider a patient holistically as comorbidities, assumed that they are correlated with the main disease, provide further information which can be used for the recognition of the disease of interest. This may facilitate the diagnostic procedure. Data-driven disease recognition approaches might support physicians in diagnosing a patient faster by simultaneously taking proper consideration of comorbidities.

In statistics, disease prediction is a typical case of classification (e. g. Dangare and Apte, 2012; Vijayarani and Dhayanand, 2015; Gárate-Escamila et al., 2020). Two concepts for classification are distinguished: single-label classification (SLC) and multi-label classification (MLC). The difference between these two approaches lies in the number of labels, i. e. the assignment of one unit of a dataset (henceforth sample) to a specific group (henceforth class). More specific, SLC methods assume a sample to exclusively belong to exactly one class; the respective label of this class is assigned to the sample. In contrast to that, MLC approaches are able to assign multiple labels to a sample; a sample can consequently belong to one class, to several classes simultaneously or even to none. Depending on which assumptions are fulfilled by the classification task at hand, one concept might be better suited.

In literature, SLC is used considerably more frequently than MLC. Still, both approaches are used in various domains, such as text categorization (e. g. Kwon and Lee, 2003; Crammer et al., 2007; Liu et al., 2017), fault detection (e. g. Dineva et al., 2019), detection of emotion in music (e. g. Turnbull et al., 2008; Trohidis et al., 2011) and in the medical context. As an example for MLC, Abdel Maksoud et al. (2019) summarize contributions to its application to diagnose several diseases via medical image classification. Further, Farooq et al. (2017) use a multi-class convolutional neural network for Alzheimer’s disease prediction. Wosiak et al. (2018) analyze the identification of comorbidities by applying different methods for MLC. They investigate methods that ignore relations between classes and those which account for them and demonstrate that the latter ones achieve higher values for the considered performance measures. In addition, Zufferey et al. (2015) use MLC to achieve precise classification of patients with chronic diseases. They consider time series health records of patients in the MIMIC-II dataset (Saeed et al., 2011).

The performance of SLC and MLC methods is compared in different research fields. To give a few examples, Sajid et al. (2023) review the use of these methods for document classification as well as arising issues. They point out the need for MLC to appropriately assign categories to research articles based on metadata or the articles’ content. In the context of drug development, Michielan et al. (2009) recommend to apply MLC instead of SLC to predict those enzymes which are involved in the metabolism of a specific substrate. The review by Wu et al. (2021) highlights the limitations of SLC as well as the advantages of MLC in the field of microbiome research, assuming that patients may suffer from several diseases simultaneously.

Despite the frequent use of SLC and MLC for disease recognition based on health records, we are not aware of a methodological review of these two approaches with respect to their conceptual foundations, practical procedures, assumed decision spaces, and strategies for handling class imbalance. Our work aims to fill this gap. Other than Zufferey et al. (2015), Wosiak et al. (2018) and Wu et al. (2021), we explore the potential added value of considering comorbidities by applying both classification approaches and comparing their predictive performance. Consequently, the novelty and added value of our contribution lies in two complementary aspects: First, a concept-driven analysis that explains and visualizes the fundamental differences between SLC and MLC in a concise yet comprehensible manner; and second, a data-driven investigation that applies both approaches to patient-derived medical data of a hospital-based department for internal medicine and rheumatology in Bielefeld, Germany, without assuming superiority of either method in advance. By doing so, we investigate whether the consideration and simultaneous recognition of comorbidities (using MLC) improves the predictability of the disease of interest compared to a recognition of the disease in isolation (using SLC).

We explain the concept of statistical classification — henceforth classification — in Section 2. In this regard, we go into more detail about SLC and MLC and highlight the differences in their decision spaces. We explain the process of model training and evaluation and introduce selected classifiers in this context. Furthermore, we discuss class imbalance in both settings. Section 3 describes a case study to recognize a disease of special interest in chronic pain patients. We describe our data and state the model specifications more precisely; further, we present and discuss the findings of the analysis. We conclude our work in Section 4 where we also provide an outlook on future research and possible extensions.

## 2 Classification

Classification is an established supervised learning concept to assign class memberships to samples based on observed variables. In the following, we describe the rationale of classification while highlighting the differences between SLC and MLC. Moreover, we introduce specific classifiers which we use in the case study in Section 3. More detailed explanations of SLC and MLC are provided in James et al. (2021) and Herrera et al. (2016), respectively.

### 2.1 Single-label and Multi-label Classification

We consider tuples (***x****_i_, **y**_i_*) which consist of covariates ***x****_i_* = (*x_i_*_1_*, x_i_*_2_*, …, x_ip_*) and target variables 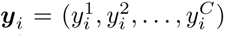, where *i* ∈ {1*, …, n*} indicates the *i^th^* sample of a dataset, *n* is the number of samples, *p* is the number of covariates, and *C* is the number of classes. The target variable is a vector ***y****_i_* ∈ {0, 1}*^C^* whose *j^th^* component is equal to one if sample *i* belongs to class *j* and zero otherwise, i. e. ∀ *j* ∈ {1, 2*, …, C*}:

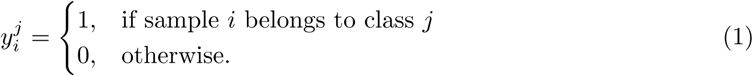

The vector ***y****_i_* represents the ground truth and is henceforth called ‘true labelset’.

Classifiers aim to suitably map observed covariates ***x****_i_* to the target; this mapping is based on a model. The classifiers are trained on true labelsets (refer to Section 2.2); the resulting predictions are given as 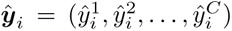. The entries of this vector are either discrete or continuous, depending on the modeler’s choice. Discrete entries are similarly defined as in Equation (1), i. e. ∀ *j* ∈ {1, 2*, …, C*}:

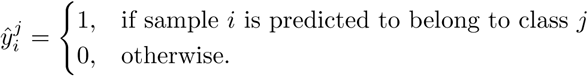

Continuous predictions can e. g. express probabilities or average votes for class memberships, with 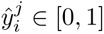 for every *j*. In the following, we refer to ***ŷ****_i_* as ‘predicted labelsets’, both for discrete and continuous predictions.

SLC and MLC differ in the fundamental assumption of how many classes a sample can belong to (ground truth). This naturally translates to a difference in the number of labels which a classifier can assign to a sample (prediction). SLC assumes that each sample belongs to exactly one of *C* classes, i. e. the classes are mutually exclusive. Consequently, ***y****_i_* is a unit vector so that 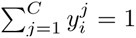 holds. The latter is also required for ***ŷ****_i_*. In discrete prediction, a classifier assigns exactly one of the *C* labels to each of the samples *i* (refer to Section 2.2). In case of continuous prediction, the vector ***ŷ****_i_* expresses the probabilities for the classes of sample *i*. A discrete predicted labelset can still be derived from a continuous one, for example by choosing the label with highest predicted probability. MLC is less restrictive than SLC on ground truth assumptions in the sense that it allows samples to belong to several classes simultaneously, including the option that a sample belongs to none or all of the considered classes. Thus, ***y****_i_* is not necessarily a unit vector but its components are equal to one for every class *j* that sample *i* is part of. Consequently, MLC potentially assigns multiple predicted labels to a sample. In case of continuous prediction, 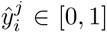 for every *j* without any further constraint on their sum.

To illustrate application scenarios, we first consider the simplest case of binary classification with *C* = 2 possible outcomes. One may, for example, aim to classify patients as either suffering from a coronary heart attack or not. Here, the considered outcomes ‘yes’ and ‘no’ are imperative for SLC; each patient can exclusively belong to one of these classes. In other contexts, however, the two considered classes do not necessarily have to be mutually exclusive. Think about patients who suffer from a coronary heart attack or a pulmonary embolism for instance. Both diseases can occur simultaneously. It depends on the modeler whether the classes are assumed to be mutually exclusive, e. g. due to the low probability of co-occurrence; in that case, one would opt for SLC. Alternatively, the samples may be assumed representative for a population in which simultaneous or no occurrences are considered relevant; this would support the use of MLC.

In both SLC and MLC, the binary case can be extended to more than two classes, i. e. to *C >* 2. Consider a set of patients who suffer from a coronary heart attack, a pulmonary embolism or asthma. There are three classes which they can belong to. As before, one may exclude co-occurrences, resulting in SLC; this is called ‘multi-class SLC’. However, it is also possible that none, several or even all of these classes apply to a patient. If we relax the assumption of mutual exclusiveness, a patient can be assigned up to three labels simultaneously, as done in multi-class MLC. In practice, the term ‘multi-class’ is commonly used in the context of SLC; consequently, the additional terms ‘single-label’ and ‘multi-class’ are often only used when particularly relevant.

The different assumptions on simultaneous class membership in SLC and MLC lead to differences in the state spaces of the labelset, the decision spaces and the classification procedures. In Figure 1, we graphically illustrate the decision and state spaces for *C* = 2 and *C* = 3 classes, both for discrete and continuous prediction and both for SLC and MLC: The state and decision spaces of the *C*-dimensional labelset appear as a square and a cube, respectively; each edge orientation represents one class, each corner a combination of class assignments from {0, 1}*^C^*. For example, the point (0, 1, 0) of the cube stands for exclusive membership to the second out of three classes; the point (0, 1, 1) represents simultaneous membership to the second and third out of three classes. While MLC allows every combination of class memberships, SLC requires 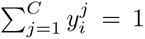. Thus, both the state and decision spaces for SLC are true subsets of those for MLC. For example, the labelsets (0, 0) and (1, 1) can only occur in the MLC setting. Assumptions on true class membership naturally extend to the set of possible predictions. Green color stands for the state space of the true labelset ***y****_i_* (all possible true labelsets), orange color marks the decision space of the predicted labelset ***ŷ****_i_* (all possible predicted labelsets). For discrete predictions, these spaces are identical. For continuous prediction, the decision space expands substantially: For *C* = 2, it stretches to the diagonal (in case of SLC) and the entire surface (MLC); for *C* = 3, it fills a triangular surface (SLC) and the entire cube (MLC).

**Figure 1:**
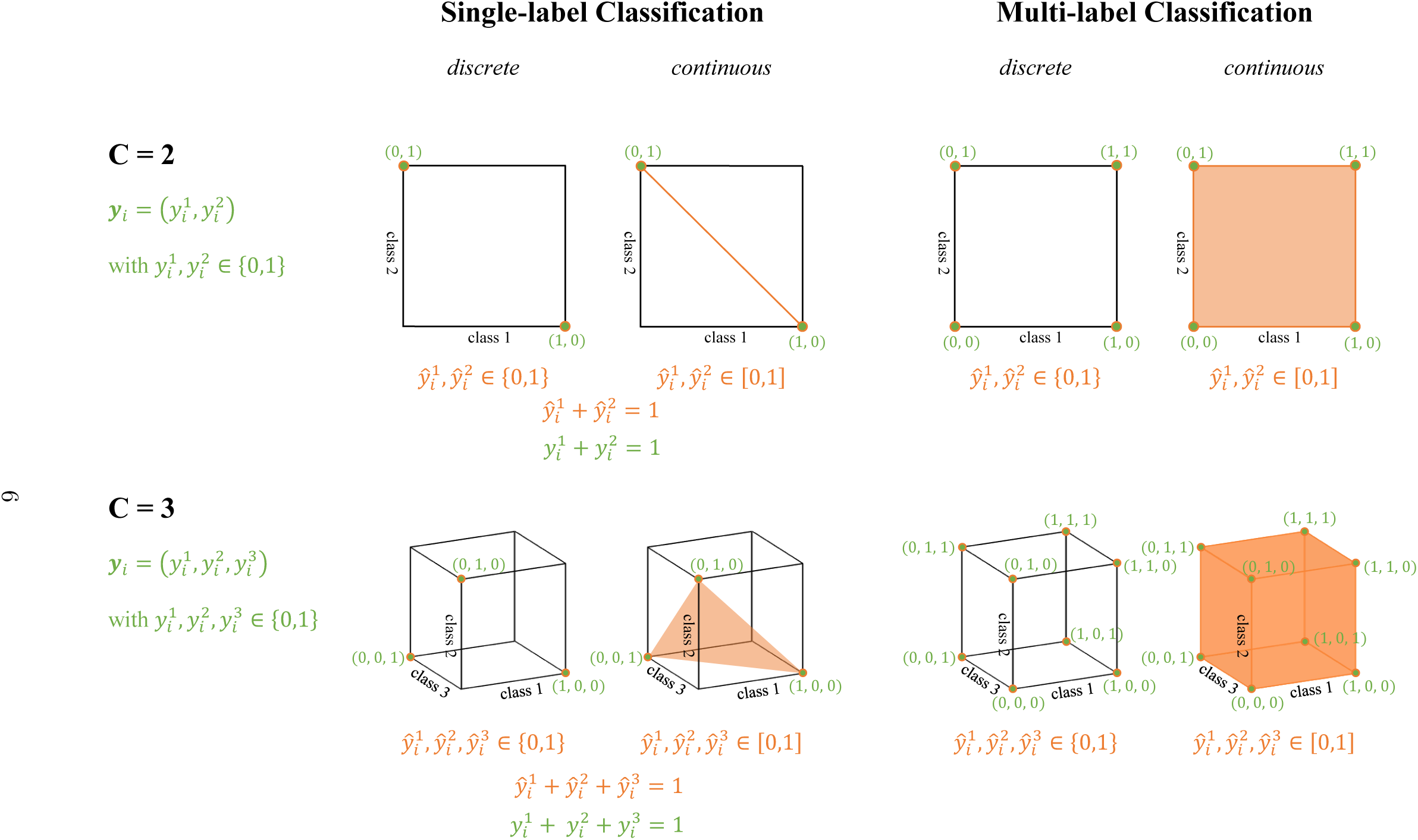
State spaces of ***y****_i_* and decision spaces of ***ŷ****_i_* for any sample *i* ∈ {1*, …, n*}. SLC and MLC are illustrated for *C* = 2 and *C* = 3 classes and for both discrete and continuous prediction. In green, potential true labelsets are depicted, whereas the possible predicted labelsets are shown in orange.

### 2.2 Model Training and Prediction

To make meaningful predictions, a classifier needs to learn how to suitably map the covariates ***x****_i_* to the true labelset ***y****_i_*. To that end, the classifier is trained on a subset of the data (henceforth training set). To assess how accurate the learned mapping classifies new data, it is applied to the remaining subset of the data (henceforth test set) which was not used for model training. By comparing the predicted labelset to the true labelset, the model performance can be evaluated using different measures, e. g. sensitivity, specificity, precision and F1-score. A perfect model, predicting the correct class for each sample, would achieve values of one in these four measures. Thus, the closer the value is to one, the better the model’s performance. Subsequently, the learned mapping can be applied to new data which is completely independent of the training and test set (so-called validation set).

A large variety of classifiers is available. In the case study in Section 3, we use tree-based methods, namely decision trees (DTs) and random forests (RFs), *k*-nearest neighbors (*k*-NN) classifiers, logistic regression models (LRMs) and neural networks (NNs), specifically multi-layer perceptrons (MLPs). The selection is motivated in Section 3. We roughly outline these methods in the following.

DTs employ step-wise separation of the covariate space into distinct regions (Hastie et al., 2009; James et al., 2021). The splitting rules are chosen such that most training samples are correctly classified; the predicted label results as the mean label of all samples falling into such a region. To build an RF, a fixed number of DTs is fitted to the data, each DT making a prediction per sample. The final prediction is then made by majority vote. Majority voting is also used for *k*-NN, considering the *k* nearest points of an observation ***x****_i_* (Hastie et al., 2009) to assign a class label. The *k* nearest and already classified neighbors are selected using a distance metric, e. g. the Euclidean distance or the Manhattan distance. An LRM is a regression model (James et al., 2021) which connects covariates to a target variable that lies in [0, 1] and represents the probability for a certain class membership. The link between the covariates and target variables is established through the logistic function which maps the real line to the unit interval [0, 1]. Accordingly, the probabilities for the different classes are modeled indirectly via the ***x****_i_*. Regarding NNs, a broadly used type are MLPs (Popescu et al., 2009). They consist of at least three layers: an input layer, one or more hidden layers and an output layer, each connected to the previous and the following one. NNs predict an outcome by transmitting the input data through all layers by following a feed-forward architecture, i. e. without any loop that allows for a repeated run-through of the layers.

The aforementioned classifiers are directly applicable in the context of SLC. For MLC, the classifiers are combined with one out of three common MLC workflows, namely (i) adaptation-based algorithms, (ii) ensemble-based methods or (iii) transformation-based methods. These are explained in detail in Herrera et al. (2016). Adaptation-based approaches are classifiers, such as a *k*-NN classifier, which are adapted from the single-label to the multi-label setting so that they are able to predict multiple labels simultaneously. Ensemble-based methods combine the results of a set of different single-label classifiers to predict a multi-label vector. Transformation-based methods handle the MLC task by transforming the problem into an SLC task, either into a binary or into a multi-class classification tasks. We apply transformation-based methods in the case study in Section 3 and therefore discuss them in more detail here.

For transformation-based methods, three main approaches can be further distinguished: binary relevance (BR), label powerset (LP) and classifier chain (CC). Figure 2 graphically visualizes these three approaches. As an example, we consider an MLC problem with *C* = 3 non-exclusive classes. The original dataset is shown at the top of the figure, the three approaches to predict the true labelset 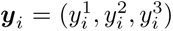 for a sample *i* are illustrated below. For them, covariates are highlighted in gray; exemplary discrete predictions are shown in white.

**Figure 2:**
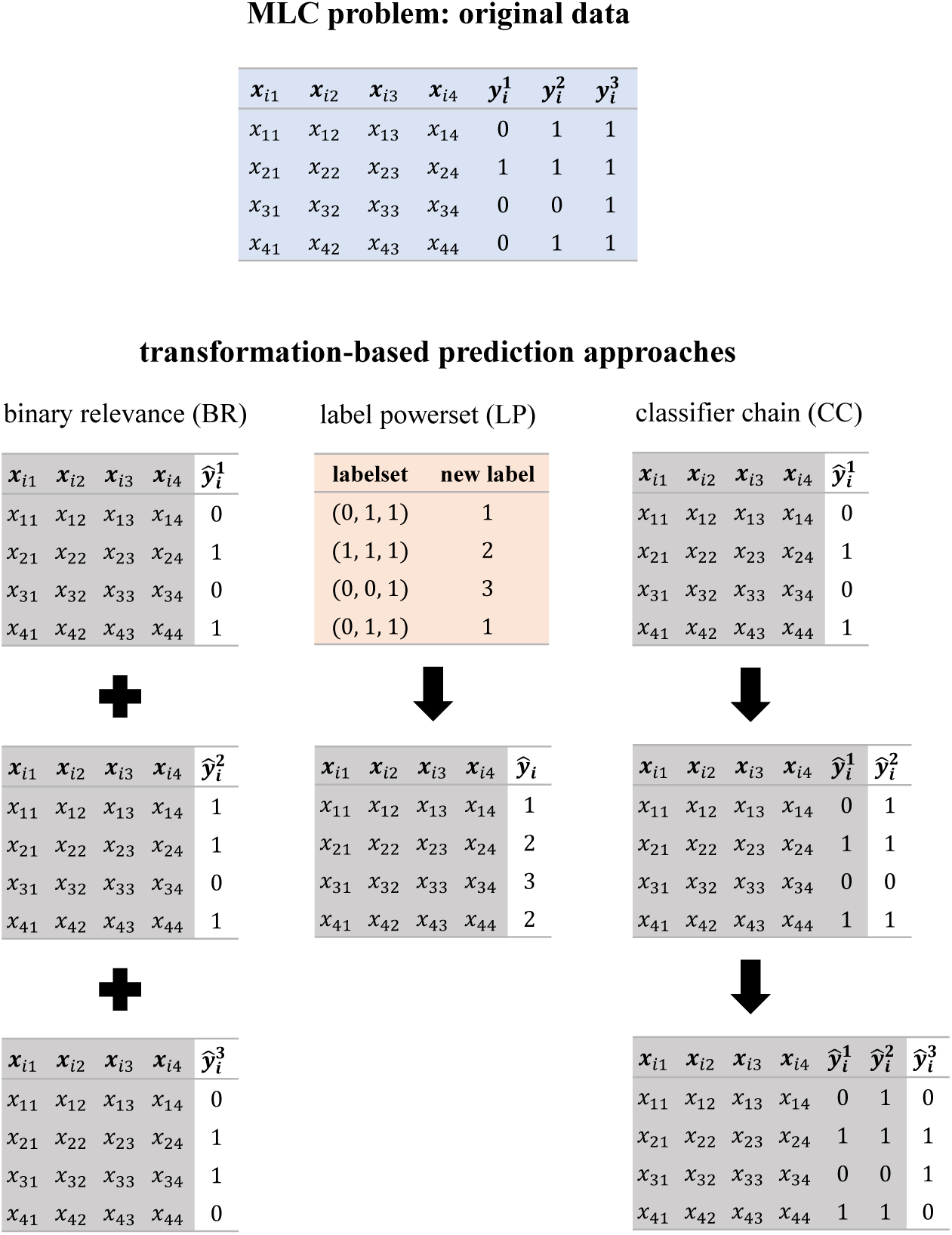
Visualization of transformation-based methods BR, LP and CC in the MLC setting to predict the target vector 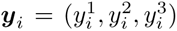. Exemplary original data is given in the blue table at the top. BR fits a binary single-label classifier for each label based on the covariates (gray shaded columns). The resulting predictions (white shaded columns) are combined (visualized by ‘+’) to full labelsets. For LP, the true labelsets are transformed into new classes (orange shaded table); based on this, the multi-label multi-class task turns into a single-label multi-class problem. For CCs, the labels are predicted consecutively. The label order for the prediction in a CC is alterable and may impact the outcome. The example shows discrete prediction; continuous prediction works the same way.

BR divides the multi-label multi-class task into *C* independent single-label binary classification tasks and merges the results afterwards (indicated by ‘+’) (Zhang et al., 2018). In our example, *C* = 3 binary single-label classifiers are used to predict *C* = 3 labels 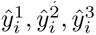 per sample *i*. The set 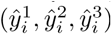 forms the predicted labelset. The approach can also be called an ensemble of binary classifiers as it combines BR with the ensemble-based approach (Herrera et al., 2016). Read et al. (2011) state that BR is often seen critically due to the fact that label correlations are unconsidered; however, they highlight the resistance to overfitting and the low computational complexity as convenient properties of BR.

The LP approach transforms the multi-label multi-class task into a single-label multi-class classification problem (Herrera et al., 2016) by increasing the number of classes: Every possible labelset is treated as a new single class, e. g. when originally dealing with *C* = 3 classes, there are 2*^C^* = 8 possible labelsets and therefore eight classes for the new single-label multi-class problem. This allows the application of well-established SLC techniques. Nonetheless, one big disadvantage of this method is the large number of classes after transformation even for small numbers of classes in the original problem. In addition, the new classes appear with no order, although their content-related closeness varies. But above all, the probability of unobserved labelsets increases with the number of classes, and it is impossible to predict unobserved labelsets using LP (Read et al., 2011).

Other than BR, a CC takes into account class correlations as it performs consecutive step-wise binary single-label prediction for the different labels (indicated by the arrows) (Read et al., 2011): CCs start by using the observations ***x****_i_* to predict the first label 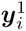 for every sample, then, it uses the observations ***x****_i_* and the predictions 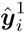, which are now considered as truth, as covariates to predict the second label 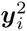. Afterwards, the observations ***x****_i_* and the predictions 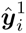 and 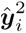 are used as covariates to predict 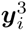. This procedure is continued until 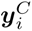 is reached. For model training, the respective elements of ***y****_i_* are used in every step of the chain. Special attention should be paid to the order of the label predictions: The order might highly affect the performance of a CC (Read et al., 2011) as incorrect predictions in early labels can lead to a bias in the following ones. The use of multiple CCs (referred to as an ensemble of classifier chains, ECC) can help to mitigate the effects of the ordering (Read et al., 2011). An ECC consists of a set of CCs. Each CC is trained on randomly selected (with replacement) samples using a random label order; each CC generates a vector of predictions ***ŷ****_i_* for each *i*. The predictions for the *i^th^* sample are subsequently averaged across all CCs and a joint decision is made for the elements of ***ŷ****_i_*.

### 2.3 Class Imbalance in SLC and MLC

Class imbalance describes the situation of markedly different class frequencies in the training set. Even if this imbalance reflects the characteristics of the underlying population, it leads to practical problems in classification: Common classifiers tend to attach greater importance to the majority class; hence, classifiers use this class as prediction for many samples to increase the number of correctly classified samples and thus achieve satisfying performance measures. Consequently, the minority class is tendentiously ignored (Guo et al., 2008). This is problematic in many applications such as fraud detection (e. g. Wei et al., 2013) or disease recognition (e. g. Cohen et al., 2006), where the minority class is of particular interest and its non-consideration might have severe consequences. This makes the issue of class imbalance a relevant and widely discussed topic (e. g. in Guo et al., 2008; Sahare and Gupta, 2012; Gosain and Sardana, 2017).

Different methods for handling class imbalance are available. Among these are random over-sampling (ROS, Fernández et al., 2018) and random under-sampling (RUS, Fernández et al., 2018) as well as synthetic data generation, e. g. via the ‘Synthetic Minority Over-sampling Technique’ (SMOTE, Chawla et al., 2002). The idea behind all these approaches is to artificially equalize class sizes, either by reducing or by increasing them. ROS randomly selects samples (with replacement) of the minority class and replicates them until the number of samples reaches the number of the majority class. As a consequence, ROS entails the risk of overfitting due to duplicates in the data (Batista et al., 2005). RUS, in contrast, randomly selects samples of the majority class and eliminates them from the dataset until the classes are of the same size. A drawback of this technique is that the dataset is reduced and potentially informative samples are removed (Batista et al., 2005). Synthetic data generation is the process of creating artificial samples for the minority class, which are based on the real data. SMOTE uses the covariates of the *k* nearest minority samples of the original dataset and interpolates between them (Chawla et al., 2002). This technique avoids the problem of overfitting (Batista et al., 2005). Nevertheless, the synthesis of new data is exclusively based on the minority class while the majority class remains unconsidered during this process. Consequently, samples that are ambiguous in terms of their class membership may be generated (Kosolwattana et al., 2023), i. e. synthesized data that should belong to the minority class can simultaneously be close to the majority class.

The described strengths and weaknesses are well known and extensively analyzed in the context of SLC. For MLC, additional difficulties arise, especially when using CCs, where the predictions of labels in the chain are based on the predictions of preceding labels. If these are heavily biased, e. g. due to imbalanced classes, the prediction of the subsequent labels might be biased as well (Senge et al., 2014). In addition, the methods described above for handling imbalanced classes are less straightforward to be applied to CCs than in the SLC setting: The true labels might be correlated, and therefore, correcting for an imbalance in one label using, e. g. RUS may cause a new imbalance in a different label. A possible way to handle this is to use a CC with RUS in such a way that RUS is integrated in each step of the CC before predicting the next label (Liu and Tsoumakas, 2020). Another approach to treat class imbalance in MLC is to consider the labels jointly, i. e. as labelsets as for LP, and to apply RUS or ROS afterwards (Charte et al., 2013). However, if the size difference between the most frequent and the rarest class is large, this procedure can lead to the dataset becoming either extremely small or extremely large.

To avoid these extrema, we apply a combination of ROS and RUS (Shamsudin et al., 2020) in the real-data case study in Section 3. The difference between this combined procedure in the context of SLC and MLC is that in SLC, ROS and RUS are applied to the *classes* whilst in MLC, ROS and RUS are applied to the *labelsets*. In the following, we refer to ROS and RUS in the context of SLC as S-ROS and S-RUS, respectively, and in the context of MLC as M-ROS and M-RUS.

## 3 Case Study: Recognition of Immune-mediated Inflammatory Diseases using SLC and MLC

‘Immune-mediated inflammatory diseases’ (IMID) is a generic term for a group of still incurable diseases with systemic (i. e. affecting a whole organ system) inflammation, for instance, rheumatoid arthritis, spondyloarthritis and others (McInnes and Gravallese, 2021). These diseases belong to a class of inflammatory autoimmune diseases which are driven by genetic and environmental factors (Anaya et al., 2006) and are caused by an improper inflammatory response of the body’s immune system to self-antigens (Rahman et al., 2010). Suffering from IMID leads to pain and other symptoms, and diminishes functioning and quality of life (Russell et al., 2011). Effective therapies which can alleviate the pain (usually consisting of basic therapeutic agents and a glucocorticoid) are already available (McInnes and Gravallese, 2021).

Fast diagnosis facilitates the contemporary initiation of a proper therapy which might reduce the complaints and avoid severe long-term consequences. An important part of the standard diagnostic procedure for IMID is measuring the value of the C-reactive protein (CRP) in the blood, as this is a valid indicator for an inflammation (Pope and Choy, 2021); CRP is often increased in IMID. A patient with an increased CRP and suspicious pain is usually referred from primary to secondary care unit to confirm IMID. However, long waiting times for an appointment delay the start of suitable therapy. Such waiting times arise due to limited capacity in the secondary care units, which is, among others, strained by many other, often unnecessary referrals. Inaccurate referrals, again, stem from the fact that a precise diagnosis is challenging for non-specialized primary care physicians. More reliable identification of IMID patients in primary care would relieve the secondary care units and improve the chances of providing rapid treatment for these patients.

IMID is frequently accompanied by comorbidities (McInnes and Gravallese, 2021), e. g. fibromyalgia syndrome (FMS), osteoarthritis (OA) and other pain-causing diseases (OPCD). This can lead to IMID being unrecognized, as the symptoms (e. g. movement restrictions or joint pain) are put on other diseases which are treated first. However, IMID is the disease among these four which needs most urgent treatment due to possible irreparable consequences. In this case study, we investigate how the recognition of IMID can be supported using statistical classification methods. In particular, we examine whether a diagnosis can be improved by a joint analysis of IMID and its comorbidities, namely FMS, OA and OPCD. In terms of statistical methodology, we ask the question whether the application of MLC constitutes an advantage over (binary and multi-class) SLC in the particular medical context. To this end, we analyze data of patients with suspected or diagnosed rheumatic diseases.

### 3.1 Data

The data analysis was done retrospectively on cross-sectional data collected by the Department of Internal Medicine and Rheumatology at Klinikum Bielefeld, Germany. All patients attended the clinic between 2019 and 2021 as they were suspected to suffer from a rheumatic disease or as they were already diagnosed with it. The original dataset contained observations from 1150 patients of 34 variables; among these were core data (age, sex, admission type, date etc.), information about glucocorticoid intake (‘prednisolone’) and the CRP value. Further, there were results of different scales to allocate a patient’s pain to a certain disease, in particular, results of the Fibromyalgia Rapid Screening Tool (FiRST) (Perrot et al., 2010), the number of tender points as well as results of the Numeric Rating Scale (NRS) (Downie et al., 1978).

During data preprocessing, we excluded variables related to the course of a disease because we considered these variables subordinate for our research question. In addition, we discarded the body mass index and the weight category which were partially inconsistent with weight and height data. Further, 81 % of both variables were missing. Also for height and weight, the large number of missing values (80 % and 77 %, respectively) was the reason for excluding these variables from our dataset. We further removed information about the examining physician, the admission date and the binary answers to the six single questions of the FiRST questionnaire (while the accumulated score was still included for analysis). The covariates used for analysis are described in more detail in Table 1. The total proportion of missing values in the covariates is 6.4 % and relates to the tender points (28 %), NRS (20 %), the duration of symptoms (10 %), CRP (4 %), the dose of prednisolone (1 %) and the intake of prednisolone (1 %) (refer to Figure A1 in the appendix). We decided to not impute missing values as health records are highly individual and the dataset is rather small regarding the number of patients as well as the number of variables. Therefore, we perform our analysis using complete data of 663 patients.

**Table 1:** Description of the covariates used for analysis.

| Variable | Description | Type | Values/ranges | Unit |
| --- | --- | --- | --- | --- |
| Age | Age of the patient | numeric | [19; 88] | years |
| Sex | Sex at birth of the patient | binary | male/female |  |
| Admission type | Admission type | binary | in-/outpatient |  |
| CRP | Measured C-reactive protein | numeric | [0.25; 149.9] | mg/l |
| Sum FiRST | Sum of positively answered questions of Fibromyalgia Rapid Screening Tool | numeric | [0; 6] | points |
| Tender points | Number of pain-sensitive pressure points on body | numeric | [0; 18] | points |
| NRS pain | Numeric rating scale to quantify the pain level | numeric | [0; 10] | points |
| Prednisolone | Prednisolone intake | binary | yes/no |  |
| Prednisolone dose | Dose of prednisolone intake | numeric | [0.0; 55.0] | mg/day |
| Duration symptoms | Duration of the symptoms that refer to the main diagnosis | numeric | [1; 600] | months |

Each patient underwent investigations for IMID, FMS, OA and OPCD; however, the dataset only contains up to three diagnoses, each assuming to contribute to the symptoms to some extent. The main disease, i. e. the disease which contributed most to the symptoms, was recorded as primary diagnosis. The diagnoses represent class assignments made by physicians. We acknowledge that they may deviate from the true underlying disease — particularly as we assume a disease not to be present if a diagnosis is missing. Still, for the purpose of this study, we assume all diagnoses to be correct and therefore constitute the ground truth. Thus, we regard the recorded data as true labelsets, and the classification task will be to derive predicted labelsets.

Based on the noted diagnoses, we derive classes considered by SLC and MLC: For binary SLC, we define the class *IMID* to contain all patients who were diagnosed with IMID as either main disease or as comorbidity. The class contains 257 patients. *noIMID* includes those 406 patients who do not fall into *IMID*. For multi-class SLC, the class *IMID* remains valid, but we moreover consider the classes *FMS\**, *OA\** and *OPCD\** which we define as the sets of all patients where the respective disease was the main disease but no IMID was diagnosed at all. The class sizes are 168, 61 and 177, respectively. *IMID*, *FMS\**, *OA\** and *OPCD\** are mutually exclusive by definition. The asymmetric consideration of IMID compared to the other diseases is motivated by the main focus of the classification task, namely revealing IMID independently of the rank of the disease. For MLC, we introduce the classes *FMS*, *OA* and *OPCD* as the sets of patients who were diagnosed with the respective disease as either main disease or comorbidity. These classes overlap with *IMID* and with each other. The class sizes are 249, 182 and 298, respectively. Table 2 provides an overview of the considered classes and the approaches in which they are used.

**Table 2:** Description of the different classes considered for SLC and MLC. The frequencies of resulting labelsets for the MLC approach are graphically provided in Figure 3 and in tabular form in Table A1 in the appendix.

| Class | Description | Approach | Class size |
| --- | --- | --- | --- |
| *IMID* | diagnosed with IMID as main disease or comorbidity | binary SLC,<br>multi-class SLC,<br>MLC | 257 |
| *noIMID* | no IMID diagnosed | binary SLC | 406 |
| *FMS\** | diagnosed with FMS as main disease, no IMID diagnosed at all | multi-class SLC | 168 |
| *OA\** | diagnosed with OA as main disease, no IMID diagnosed at all | multi-class SLC | 61 |
| *OPCD\** | diagnosed with OPCD as main disease, no IMID diagnosed at all | multi-class SLC | 177 |
| *FMS* | diagnosed with FMS as main disease or comorbidity | MLC | 249 |
| *OA* | diagnosed with OA as main disease or comorbidity | MLC | 182 |
| *OPCD* | diagnosed with OPCD as main disease or comorbidity | MLC | 298 |

### 3.2 Methodical Procedure

We investigate whether the recognition of IMID can be improved by considering comorbidities, even if these are unknown during prediction. In line with the methodological focus of our work, this means that we examine whether MLC recognizes IMID more reliably than SLC. There may be statistical methods apart from the approaches chosen here, especially apart from classification, that might exploit the specific dataset better. However, these would go beyond the scope of our research question of method comparison, so we will limit our analysis to the SLC and MLC methods presented in Section 2.

We assume MLC being advantageous over SLC especially if the disease occurrences in our data are associated with each other. Therefore, we first examine the association by performing Pearson’s *χ*^2^-test with Yates’ continuity correction (Yates, 1934). To adjust the p-values, we make use of the Bonferroni correction (Bonferroni, 1935). According to this, we observe an association between IMID and FMS, IMID and OPCD (consisting of the disease groups ‘arthralgias of unclear assignment’ and ‘others’), FMS and OPCD as well as OA and OPCD (refer to the respective (adjusted) p-values in Table A2 in the appendix).

We tackle the classification of *IMID* through the following six approaches:

- **Approach 1a: Binary SLC without rebalancing** We consider the two exclusive classes *IMID* and *noIMID* via SLC. In this basic approach, we do not account for class imbalance.
- **Approach 1b: Binary SLC with S-ROS and S-RUS** The class imbalance between *IMID* and *noIMID* in Approach 1a motivates class imbalance correction. Here, we apply a combination of S-ROS and S-RUS. We aim at the average number of samples of both classes in the training set per class as a compromise.
- **Approach 2a: Multi-class SLC without rebalancing** We consider the four exclusive classes *IMID*, *FMS\**, *OA\** and *OPCD\** and tackle the task using multi-class SLC.
- **Approach 2b: Multi-class SLC with S-ROS and S-RUS** The four classes from Approach 2a are highly imbalanced. Consequently, as in Approach 1b, we apply a combination of S-ROS and S-RUS to the training set to account for the class imbalance. Per class, we aim at the average number of samples of all four classes.
- **Approach 3a: Multi-class MLC without rebalancing** As in Approaches 2a and 2b, we consider all four diseases simultaneously. However, we now employ the non-exclusive classes *IMID*, *FMS*, *OA* and *OPCD*. In other words, we allow patients to have multiple labels simultaneously and consequently perform MLC. Figure 3 and Table A1 display the number of occurrences of true labelsets. To handle the classification task, we use the transformation-based CCs (refer to Section 2.2). BR is discarded because of its inability to consider label correlations. Further, we abstain from using LP as it is unsuited to predict unobserved labelsets (Read et al., 2011). In contrast, CCs allow us to investigate whether the prediction of FMS, OA and OPCD improves the recognition of IMID from the present data. Although only 14 out of 16 labelsets are represented in the data (ground truth), all labelsets can be predicted due to the step-wise binary prediction of the four labels using CCs. Within the CC approach, we consider different label orderings. However, as our research question implies that IMID should be predicted as last element of the target vector, we estimate the MLC models assessing only six different label orderings instead of 24. In Section 3.3, we report and discuss the results of only that ordering which leads to the highest average F1-score for the majority of all applied classifiers; that is the ordering where 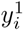 corresponds to FMS, 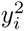 to OPCD, 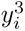 to OA and 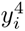 to IMID. The results of the other label orderings differ only slightly.
- **Approach 3b: Multi-class MLC with M-ROS and M-RUS** We perform MLC with the same classes as in Approach 3a, this time with a combined procedure of M-ROS and M-RUS to account for class imbalance. In contrast to the proceeding in the context of SLC (S-ROS and S-RUS, Approach 2b), we apply the rebalancing method to the true labelsets rather than to the classes (which were defined differently, and thus the procedures are not transferable to each other). The smallest class among the 14 present labelsets is the group of patients who simultaneously suffer from OPCD, OA and IMID and consists of 13 samples. The largest class contains 160 samples; these patients exclusively suffer from OPCD. Using a combination of M-ROS and M-RUS, we aim at the average number of samples of all true labelsets in the training set.

**Figure 3:**
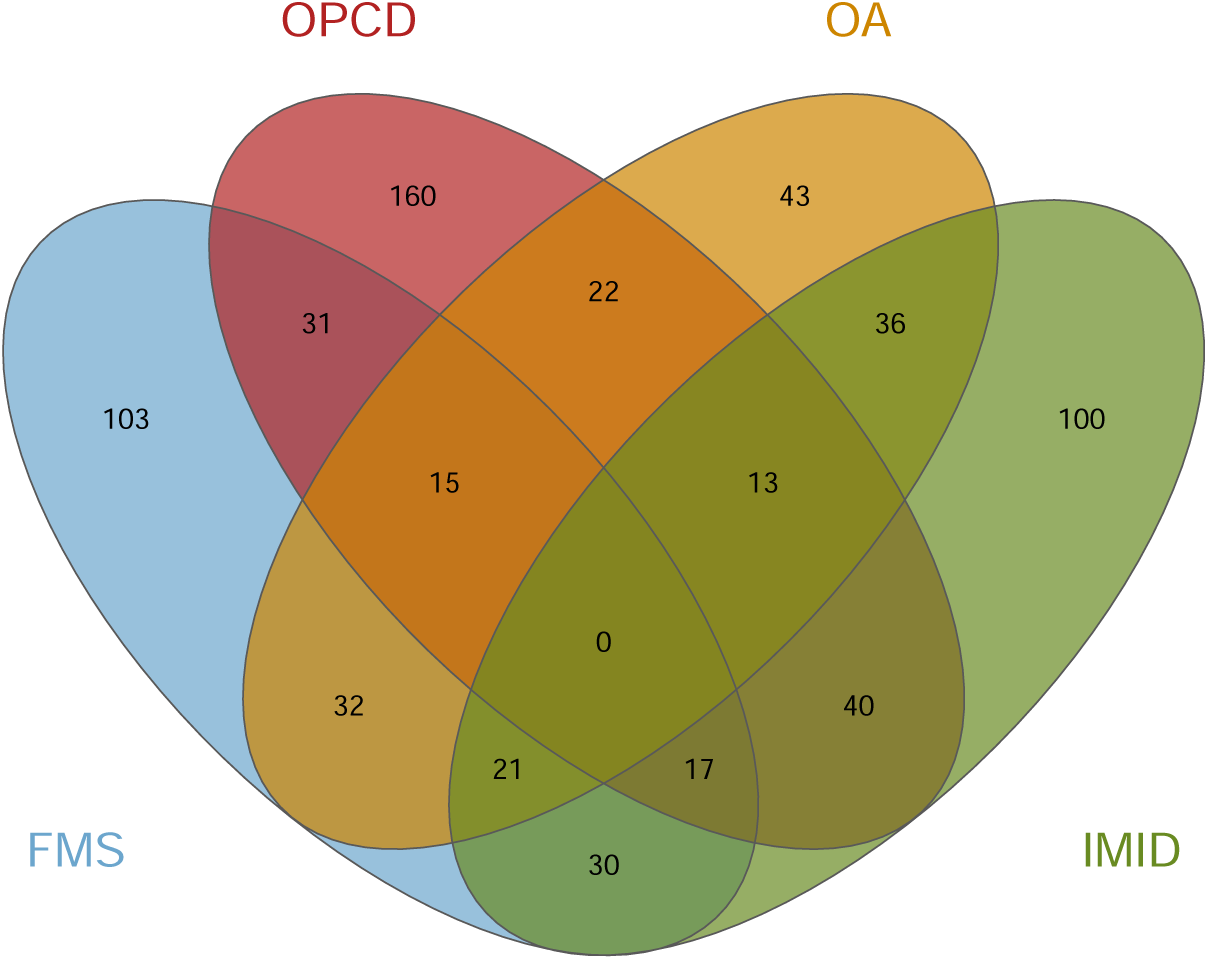
Number of (joint) occurrences of the four diseases in the dataset visualized as Venn diagram. Each patient suffers from at least one disease; no patient suffers from all four diseases. Consequently, the true labelsets (0, 0, 0, 0) and (1, 1, 1, 1) are not present in the data. A tabular representation of all numbers is additionally provided in Table A1 in the appendix.

For each approach, we apply DT, RF, LRM, *k*-NN and MLP as classifiers in order to take various requirements into account. DT, *k*-NN and LRM are easily interpretable, which is an important criterion for the use of statistical models in the medical field. The rather complex structure of medical data due to the individuality of patients, however, might require more complex models. Hence, we use RF and MLP as classifiers and accept their reduced interpretability. To account for the rather small amount of data and thus avoid overfitting of the models, we apply nested cross-validation (CV). Hence, we validate the models’ performance on different training and test splits. The outer loop is a ten-fold CV, the inner loop is a five-fold CV to optimize the hyperparameters of the classification models using a grid search. Hence, previously indicated class sizes refer to the entire dataset before splitting it into training and test data. We use macro precision, macro sensitivity as well as macro F1-score to evaluate each of the cross-validated models. We refit the optimized model using the macro F1-score, as it is the harmonic mean of the sensitivity and the precision. Due to the need for a rapid initiation of a treatment in case of the IMID, the sensitivity and the precision are of higher priority than the specificity. For the DTs, we tune the criterion to measure the quality of a split, the maximum depth of the tree and the minimum number of samples in a node to perform a further split. In addition to these hyperparameters, we optimize the number of trees for the RFs. For the LRMs, we tune the penalty norm, the tolerance for the stopping criteria as well as the used algorithm for solving the optimization problem. When estimating the MLPs, we optimize the number of hidden layers each consisting of 100 neurons, the activation function of the hidden layer, the solver, the strength of the *L*2 regularization term and the learning rate. In case of the *k*-NN classifier, we tune the number of considered neighbors *k*. Recall Figure 3 for a visualization of the number of occurrences of the four diseases in the dataset and their intersections. A tabular form of these numbers is additionally provided in Table A1 in the appendix.

### 3.3 Results and Discussion

In the following, we outline the results of the application of all models described in Section 3.2 to the data from Section 3.1. The average performance per approach and per classifier across all ten CV folds is shown in Table 3.

**Table 3:**
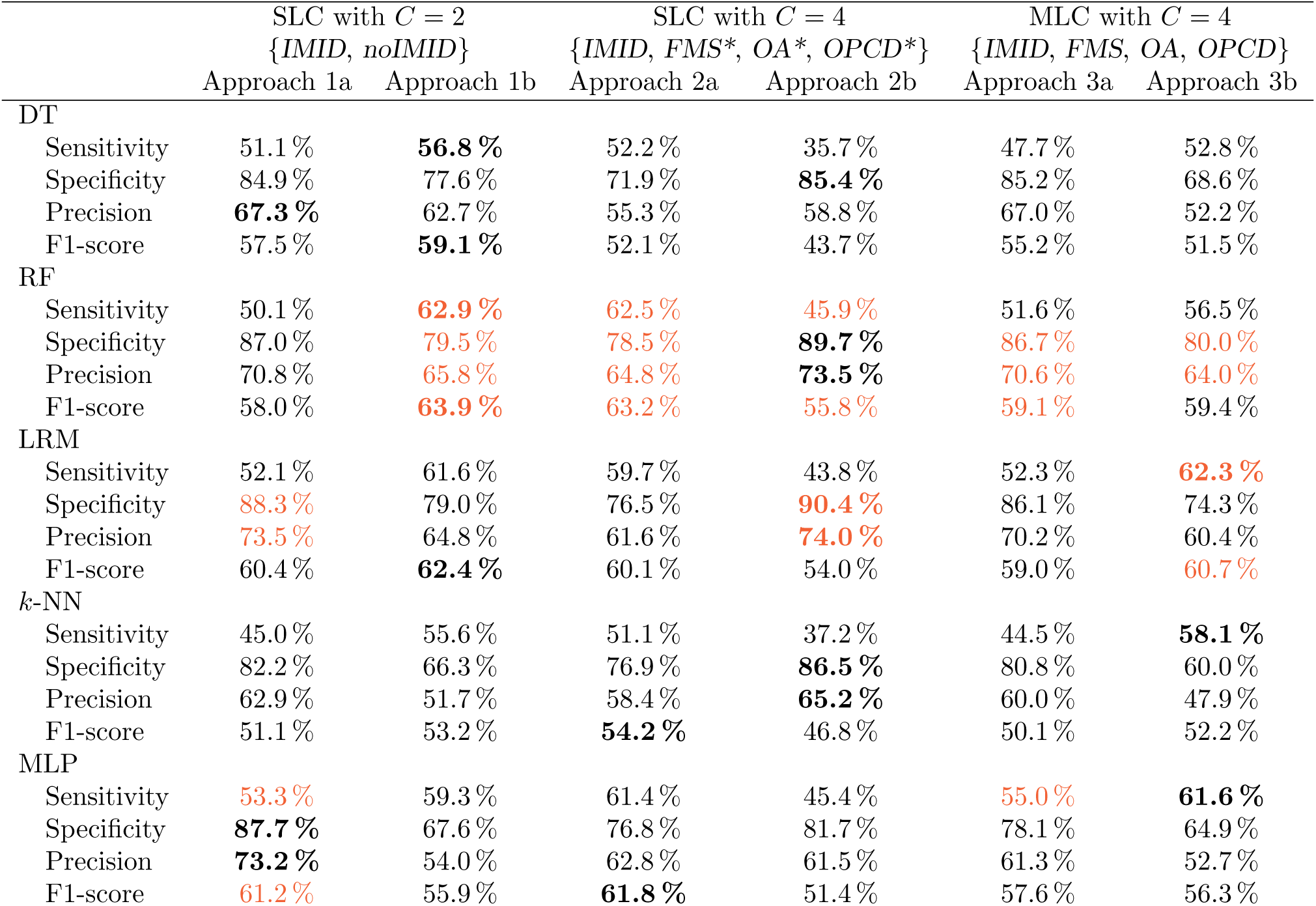
Average performance (across ten CV folds) of the six described approaches for recognizing IMID. The approach as well as the considered classes are provided in the column names. The highest average measures per approach (i. e. per column) are highlighted in orange, the highest average measure per classifier (i. e. per row; DT, RF, LRM, *k*-NN, MLP) is written in bold.

#### 3.3.1 Comparison of Classifiers

Firstly, we compare the classifiers DT, RF, LRM, *k*-NN and MLP with each other by looking at the approaches individually. The best measures (i. e. the best specificity, sensitivity, precision and F1-score) for each approach (that is, column-wise) across all classifiers are highlighted in orange.

For **Approach 1a**, the four best measures are spread out across two classifiers, namely LRM and MLP. MLP performs best regarding the sensitivity (53.3 %) and the F1-score (61.2 %), whereas LRM achieves the highest precision (73.5 %) and the highest specificity (88.3 %). The two latter values are good results; however, the remaining two measures are too low for a reliable classification. The results are quite similar for all classifiers; *k*-NN achieves the lowest value in all four measures.

In **Approach 1b**, RF performs best regarding all measures: It results in a sensitivity of 62.9 %, a specificity of 79.5 %, a precision of 65.8 % and an F1-score of 63.9 %. All measures are moderately high, but still too low for a valid use in the medical field.

The highest results of **Approach 2a** are almost identical to those of Approach 1b: RF achieves a sensitivity of 62.5 %, a specificity of 78.5 %, a precision of 64.8 % and an F1-score of 63.2 %. MLP and LRM achieve comparable results, DT and *k*-NN perform worse.

**Approach 2b** leads to a low sensitivity (at most 45.9 %, achieved by RF) as well as to a low F1-score (at most 55.8 %, achieved by RF), but a high specificity (90.4 %, achieved by LRM) and a moderately high precision (74.0 %, achieved by LRM); this pattern is recognizable for RF and LRM, whereas DT, MLP and *k*-NN result in low measures except from the specificity.

**Approach 3a** results in a low sensitivity (55.0 %, achieved by MLP) and a low F1-score (59.1 %, achieved by RF). The precision is moderately high at 70.6 %; the specificity, in contrast, is high at 86.7 % (both achieved by RF).

As in Approaches 1b and 2a, the models of **Approach 3b** show a moderate performance; viewed in isolation, almost all classifiers perform rather weak regarding the sensitivity. LRM results in the highest sensitivity of 62.3 %; the highest F1-score of 60.7 % achieved by LRM is rather low. Even the specificity of 80.0 % achieved by RF is, compared to other approaches, a weak result. Across all classifiers, the precision is rather low; it ranges from 47.9 % achieved by *k*-NN to 64.0 % achieved by RF.

To draw an interim conclusion: The best performances of the six approaches is spread out across RF, LRM and MLP, with a noticeable accumulation at RT. In contrast, DT and *k*-NN show comparably low performance. Measured by the F1-score, the RF seems to be the best suited classifier for our analysis in the majority of approaches.

#### 3.3.2 Comparison of Approaches

When comparing the six Approaches 1a to 3b with each other, we take particular account of the values written in bold in Table 3. These mark the highest average measure per classifier (that is, row-wise). Approaches 1b and 2b achieve the best results: Most of the bold-printed values are distributed between those two. In the following, we go into more detail about the differences between the six approaches.

##### Approaches with vs. without rebalancing

We first compare the approaches in pairs, namely those accounting for the same number of classes and allowing for the same number of labels, but one with and one without rebalancing (that is: a vs. b). After this, we highlight the differences between the three approaches without rebalancing (a), followed by a comparison of those accounting for class imbalance (b). Due to the importance of detecting patients suffering from IMID as well as being precise in the recognition, we pay special attention to the F1-score for the purpose of selecting the best performing approach.

Compared to the results of **Approach 1a**, the F1-score of all classifiers, except from MLP, in **Approach 1b** increased by 1.6 to 5.9 percentage points (pp); the F1-score of MLP is decreased by 5.3 pp. For most classifiers, the sensitivity increases to a considerable extent (by max. 12.8 pp), but at the same time the specificity and the precision decrease (by max. 20.1 pp and 19.2 pp, respectively). The difference between these two approaches lies in the handling of class imbalance. While the classes in Approach 1a are highly imbalanced (the entire dataset contains 257 patients with IMID and 406 patients without), both classes are equally represented in Approach 1b. Considering this class imbalance, the low sensitivity and high specificity of the models in Approach 1a are reasonable, as the positive samples (i. e. the patients suffering from IMID) are underrepresented. Consequently, the changes in results meet our expectations of an increased sensitivity and a decreased specificity. Albeit an increase in sensitivity is desirable (the models get better at predicting patients who require rapid treatment as they suffer from IMID), a decrease in specificity might lead to an overstrain of clinics resulting in long waiting times which increase the time to first treatment. Nevertheless, with regard to the F1-score, Approach 1b mostly outperforms Approach 1a.

**Approaches 2a and 2b** consider *C* = 4 classes each. Comparing these two approaches, Approach 2b results in a considerably worse sensitivity (decreased by up to 16.6 pp) but in a strongly increased specificity (by max. 13.9 pp) for all classifiers. This is similar for the precision and the F1-score: The F1-score decreases (my max. 10.4 pp), while the precision increases (by max. 12.4 pp). Only for MLP, the precision decreases by 1.3 pp. In the entire dataset, the difference of 196 samples between the largest (IMID) and the smallest class (OA) is large. Accordingly, the decrease in sensitivity and the increase in specificity were to be expected, as IMID, the class of main interest, is downsampled for the purpose of class rebalancing which reduces the amount of potentially useful information. Nevertheless, without handling class imbalance, our model might be biased and would consequently result in too many false positively predicted samples. The rebalancing, however, leads to a strong decrease in sensitivity which results in a non-identification of patients who need rapid treatment. When selecting an approach based on the F1-score, Approach 2a outperforms Approach 2b.

In **Approaches 3a and 3b**, we utilize the fact that FMS, OA and/or OPCD frequently accompany IMID; we there analyze the data using MLC. Rebalancing in the context of these models increases the sensitivity by 4.9 to 13.6 pp, but lowers the specificity by 6.7 to 20.8 pp. At the same time, the precision decreases by up to 14.8 pp. The F1-score yields an ambiguous picture, ranging from a decreases by at max. 3.7 pp to an increase by up to 2.1 pp. Similar to the difference between Approaches 1a and 1b, the increased sensitivity allows for a better recognition of those patients suffering from IMID; however, the specificity is too low to certainly identify patients without IMID. Nevertheless, Approach 3b outperforms Approach 3a based on the F1-score.

As an interim conclusion, we summarize that the difference in model performance between unbalanced and balanced approaches depends on (i) the number of considered classes and (ii) the consideration as either SLC or MLC problem. For *C* = 2 (Approaches 1a and 1b), SLC performs better at predicting IMID after rebalancing, although the sensitivity is insufficient for a disease that requires rapid treatment. For *C* = 4, however, rebalancing worsens the recognition of IMID for SLC (Approaches 2a and 2b) but improves it for MLC (Approaches 3a and 3b). This makes sense as IMID class sizes vary across approaches; consequently, it is sometimes upsampled, sometimes downsampled, resulting in varying sensitivity.

In the following, we compare the three approaches (i. e. (1a/b) binary SLC, (2a/b) multi-class SLC, and (3a/b) multi-class MLC) with each other, first without rebalancing, second with rebalancing.

##### Approaches without rebalancing

Approaches 1a, 2a and 3a differ in the aforementioned two main points: (i) the number of considered classes and (ii) the consideration as either SLC or MLC problem. Regarding (i), Approach 1a considers *C* = 2 classes, whereas Approaches 2a and 3a both distinguish *C* = 4 classes. Regarding (ii), Approaches 1a and 2a use SLC, whereas Approach 3a uses MLC. MLC allows a more generous assignment of IMID labels, since the occurrence of other diseases does not exclude this. In addition, considering comorbidities for the recognition of IMID increases the set of possibly informative covariates compared to SLC. Still, the sensitivity of all models belonging to Approach 3a are worse than the ones of Approach 2a; even compared to Approach 1a, the sensitivity achieved by DT and *k*-NN is lower. In case of RF, for instance, the sensitivity is 10.9 pp lower than in Approach 2a and just 1.5 pp higher than in Approach 1a. A possible explanation for the decreased sensitivity in Approach 3a is that the prediction of IMID is based on the prediction of the other three labels due to the construction of CCs. However, the precedent predictions might be imprecise due to highly imbalanced classes. As a result, the prediction of IMID might be imprecise as well.

##### Approaches with rebalancing

As motivated above, handling class imbalance is necessary to aim at precise models, which is why we applied rebalancing methods to our data. Among the models with rebalancing (Approaches 1b, 2b and 3b), the poor sensitivity of Approach 2b is immediately apparent. Compared to the Approaches 1b and 3b, it is 10.6 to 20.9 pp lower. The specificity, however, is highest in Approach 2b for every classifier. This also applies to the precision, except for DT. Regarding the F1-score, Approach 1b performs best in case of DT, RF, LRM and *k*-NN; in case of MLP, Approach 3b is best. Based on these results and assuming the F1-score being the most important measure, Approach 1b seems to perform best on our data. As for the unbalanced approaches, the MLC methods do not generally outperform SLC although IMID is frequently accompanied by FMS, OA and/or OPCD, and although Pearson’s *χ*^2^-test indicated an association between the diseases (refer to Table A2 in the appendix), except for IMID and OA as well as for FMS and OA. As an additional analysis, we applied MLC without consideration of the class OA, but this lead to no noticeable difference to the reported models (refer to Table A3 in the appendix).

#### 3.3.3 Contextualization of the Results

Our results present an ambiguous picture: There is no clear evidence that analyzing IMID in isolation (Approach 1a/b) leads to improved diagnostic performance when compared with a more differentiated consideration of diseases (Approach 2a/b) or the inclusion of comorbidities (Approach 3a/b). While the overall best sensitivity and F1-score is achieved by Approach 1b (62.9 % and 63.9 %, respectively), the overall best specificity and precision is reached by Approach 2b (90.4 % and 74.0 %, respectively). However, performance varies across approaches, application of rebalancing and classifiers; the only method that never outperforms the others (for no classifier and none of the four measures) is Approach 3a, i. e. MLC without rebalancing.

Weak performance or the fact that taking comorbidities into account does not lead to a clear improvement may be due to the following factors:

- **Correlation strength:** The association between the diseases may be too weak for an added value of considering comorbidities. Even if disease occurrence is correlated, this may be insufficient to provide relevant information for the recognition of IMID.
- **Missing information:** The data may miss relevant covariates for reliable recognition of IMID. It contains 13 variables, but most of them are rather related to FMS. This applies, in particular, to the FiRST questionnaire and the tender points. The CRP value is a valid indicator for any inflammation in the body; consequently, the variable is not necessarily sufficient to clearly separate the diseases from one another. For OA in particular, no typical variables (e. g., X-ray findings or other variables which query frequently occurring symptoms) are included in the dataset. This makes the recognition of comorbidities and thus the recognition of IMID more difficult.
- **Class imbalance:** The disparity in class sizes is partly substantial. This imbalance may bias the results: Our class imbalance correction often improves the classifiers’ performance, but the dataset is strongly manipulated by excluding and duplicating samples.
- **Selection of patients:** Some of the patients in the dataset were non-recently diagnosed and already received treatment before data collection. This affects, for example, the CRP value so that it no longer corresponds to the value without treatment; this makes clear pattern recognition more difficult.

Despite the inconclusive picture and the limitations of the dataset, the data application illustrates the influence that different numbers of classes and the interpretation as an SLC or MLC problem can have. It also shows how the correction for class imbalance affects the results. In doing so, it demonstrates the challenges of disease classification using a real-world dataset.

## 4 Conclusion

In this work, we provided a methodological examination of SLC and MLC approaches, with a particular focus on disease recognition based on health records. By explaining and comparing both frameworks conceptually and empirically, we illustrated how underlying assumptions, learning structures, and strategies for handling class imbalance shape predictive performance. Using patient-derived data from a clinical setting, we demonstrated the practical implications of these methodological choices, particularly regarding the simultaneous consideration of comorbidities. In doing so, our study connects conceptual clarification with empirical validation, thereby contributing to a more systematic understanding of when and how SLC or MLC may be advantageous in medical classification tasks.

In a case study, we investigated whether the simultaneous consideration of comorbidities results in a more reliable recognition of pain-causing diseases. The aim of the analyses was to compare SLC and MLC, together with other choices in connection with the training and prediction procedure, rather than to identify the best possible model for recognizing the disease of interest. In detail, we performed a case study on 663 patients with suspected or diagnosed rheumatic diseases, namely IMID, FMS, OA and OPCD. The study investigated the effect of considering different numbers of classes, and of interpreting the problem as SLC or MLC. In the context of MLC, we chose CC, a transformation-based method, in order to account for label correlations. For both SLC and MLC, class imbalance was handled using a combination of ROS and RUS. To examine the impact of these procedural decisions, we applied six different approaches to the same data. Each of them used DTs, RFs, LRMs, *k*-NN and MLP as classifiers.

The analyses yielded an ambiguous picture. Each method choice entailed different strengths and weaknesses: between SLC or MLC, between the number of classes, the correction for class imbalance, the classification method, and the evaluation measure. This underlines the importance of addressing each individual problem in an equally individual manner, especially when the implications can be significant, such as in patient referrals: How strongly are interdependencies between diseases assumed? How complex (or, e. g., non-linear) does the relationship between disease and covariates appear, and to what type of statistical classification method does this lead? Which aspect of performance do we consider most important, e. g., sensitivity or specificity?

A look at the situation underlying the examined use case helps to, for example, contextualize the relevance of different performance measures: Current practice shows that (too) many suspected cases are referred to rheumatology departments, where the diagnosis is often not confirmed by specialists. This unnecessarily prolongs waiting times for patients who are actually affected by IMID. At the same time, it remains crucial that every patient has the opportunity to be examined in a specialized clinic.

This leads to the highly relevant point that computational prediction tools should always serve as decision *support* for treating physicians: complementing, but never replacing, their clinical judgment. Against this background, the question arises of how classification results should be presented. In our use case, we applied hard (discrete) classification thresholds, predicting patients as either diseased or non-diseased, as this approach suited the research questions of comparing performance between methods. In the context of decision support, however, it is immediately possible to generate continuous predictions, such as probabilities. Each of the approaches discussed here is capable of providing such information.

Independent of the specific use case, the analyzed data, and the observed performance results, it is important to consider the conceptual advantages of MLC compared to SLC. While SLC requires a decision for a single class, even when the available covariates may indicate multiple possible diseases, MLC allows for the simultaneous assignment of several conditions to one patient. This reflects a more realistic representation of clinical practice, where multiple diseases can coexist and should therefore be jointly considered in diagnostic support. At the same time, it is important to emphasize that neither approach conceptually dominates the other: In practice, models with a larger number of classes do not necessarily yield better results than those with fewer classes, and MLC is not inherently superior to SLC. In practical implementation, computational costs and the effort required for correcting class imbalance —both of which tend to increase with a higher number of classes and when using MLC — also need to be carefully weighed.

In summary, our study highlights the potential of statistical decision support to improve clinical decision-making. The underlying methods extend well beyond the health context. Their usefulness increases with the availability of high-quality data and the careful selection of appropriate statistical approaches: The better the alignment between the use case, the data, and the methodological assumptions, the more reliable and practically relevant the results become. Future research should therefore focus on refining data quality, exploring advanced modeling techniques, and developing transparent, interpretable tools that can effectively assist physicians — and professionals in other domains — in making informed decisions.

## Data Availability

Due to data privacy protection, we are not allowed to share the original data.

## Glossary

BR: binary relevance
CC: classifier chain
CRP: C-reactive protein
CV: cross-validation
DT: decision tree
ECC: ensemble of classifier chains
FiRST: Fibromyalgia Rapid Screening Tool
FMS: fibromyalgia syndrome
IMID: immune-mediated inflammatory disease
LP: label powerset
LRM: logistic regression model
*k*-NN: *k*-nearest neighbors
MLC: multi-label classification
MLP: multi-layer perceptron
NN: neural network
NRS: Numeric Rating Scale
OA: osteoarthritis
OPCD: other pain-causing diseases
RF: random forest
ROS: random over-sampling
RUS: random under-sampling
SLC: single-label classification
SMOTE: Synthetic Minority Over-sampling Technique

## Author Contributions

Conceptualization: Sophie Schmiegel, Christiane Fuchs, Hannah Marchi

Data curation: Martin Rudwaleit, Marvin-Hendrik Röchter

Formal analysis: Sophie Schmiegel

Funding acquisition: Christiane Fuchs, Martin Rudwaleit

Supervision: Christiane Fuchs

Visualization: Sophie Schmiegel

Writing – original draft: Sophie Schmiegel

Writing – review and editing: Sophie Schmiegel, Christiane Fuchs, Hannah Marchi, Martin Rudwaleit, Marvin-Hendrik Röchter

## Ethics Statement and Consent to Participate

Ethical approval was given by the Ethics Committee of the Westphalia-Lippe Medical Association and the University of Münster (Ethik-Kommission der Ärztekammer Westfalen-Lippe und der Westfälischen Wilhelms-Universität Münster); case number: 2021-426-f-S. All patients agreed on participating in this study.

## Computational Details and Data Availability Statement

The data preprocessing was performed using R version 4.4.2 (R Core Team, 2024). The SLC models as well as the MLC models were fitted using Python version 3.11.9 (Python Software Foundation, 2024). The code used for model fitting is available at https://github.com/fuchslab/SLC_MLC_Disease_Recognition.

Due to data privacy protection, we are not allowed to share the original data.

## Acknowledgements

We thank Tamara Schamberger, Philipp Hege and Valentina Lakusta for helpful comments and discussions.

## A Appendix

### A.1 Case Study: Overview of Missing Values in the Data

**Figure A1:**
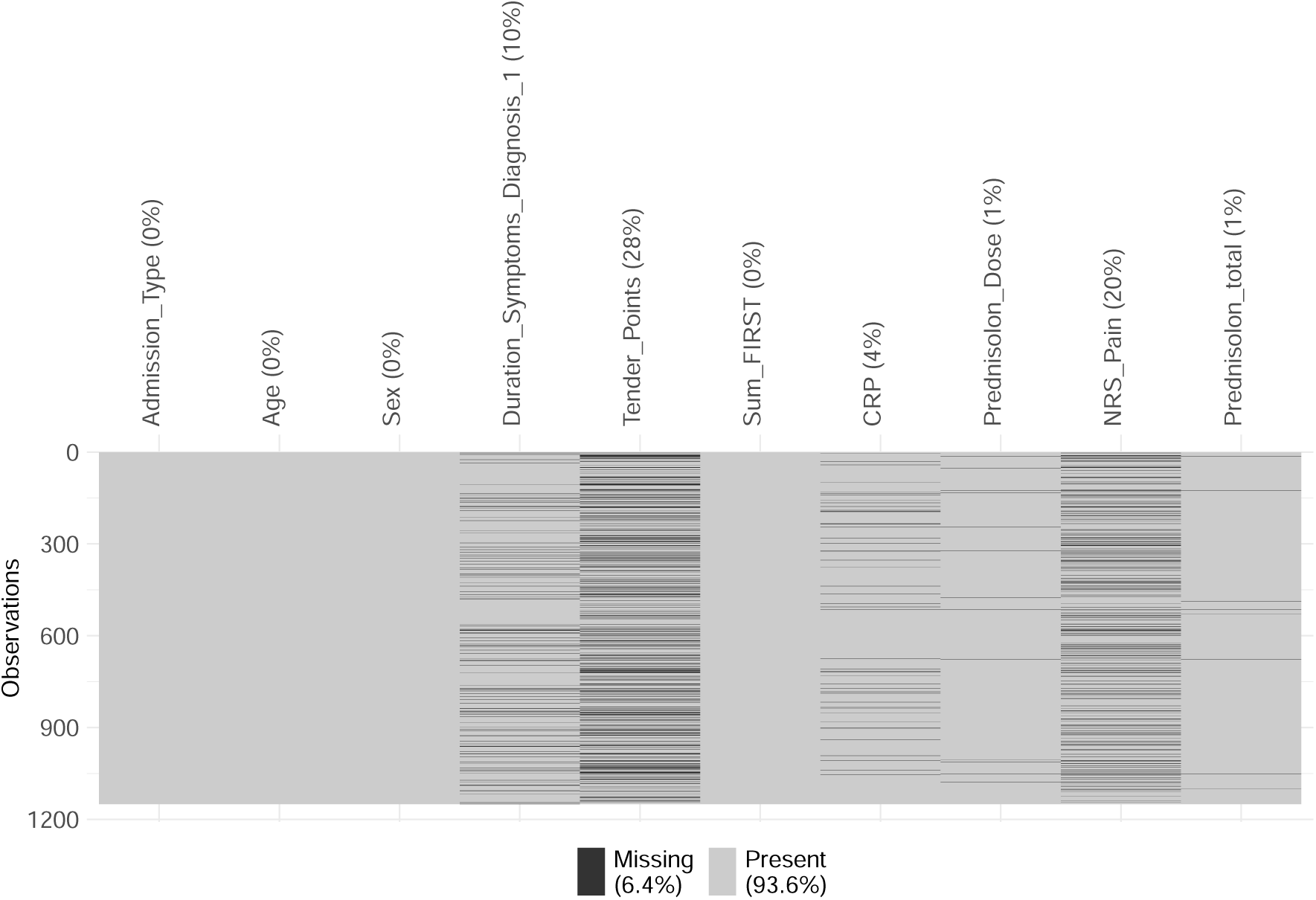
Frequencies of missing values in those variables that were used for statistical analysis. The set of 1150 patients was reduced to the complete cases, resulting in 663 patients.

### A.2 Case Study: Additional Material on the Results

#### A.2.1 Counts of True Labelsets

**Table A1:**
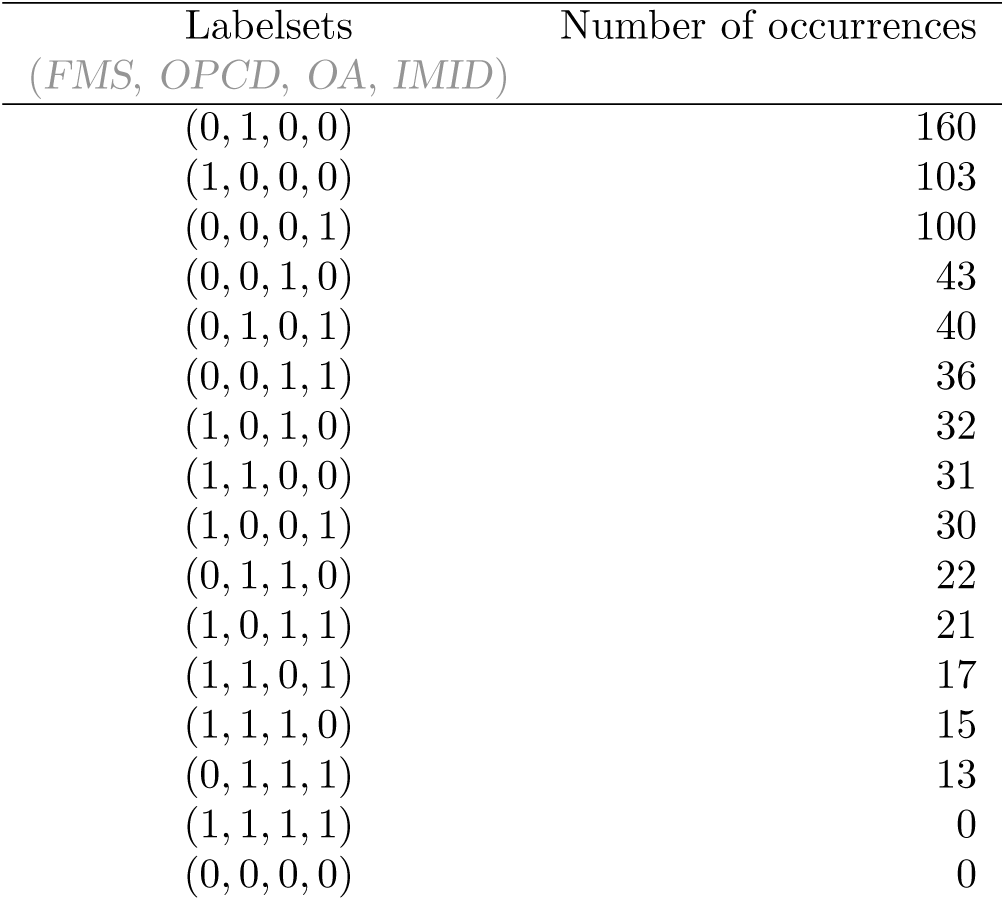
True labelsets and their numbers of occurrence in the dataset under consideration. 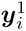 corresponds to FMS, 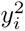 to OPCD, 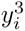 to OA and 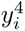 to IMID.

| Labelsets<br>(*FMS*, *OPCD*, *OA*, *IMID*) | Number of occurrences |
| --- | --- |
| (0, 1, 0, 0) | 160 |
| (1, 0, 0, 0) | 103 |
| (0, 0, 0, 1) | 100 |
| (0, 0, 1, 0) | 43 |
| (0, 1, 0, 1) | 40 |
| (0, 0, 1, 1) | 36 |
| (1, 0, 1, 0) | 32 |
| (1, 1, 0, 0) | 31 |
| (1, 0, 0, 1) | 30 |
| (0, 1, 1, 0) | 22 |
| (1, 0, 1, 1) | 21 |
| (1, 1, 0, 1) | 17 |
| (1, 1, 1, 0) | 15 |
| (0, 1, 1, 1) | 13 |
| (1, 1, 1, 1) | 0 |
| (0, 0, 0, 0) | 0 |

#### A.2.2 Pearson’s ***χ*^2^**-Test

**Table A2:**
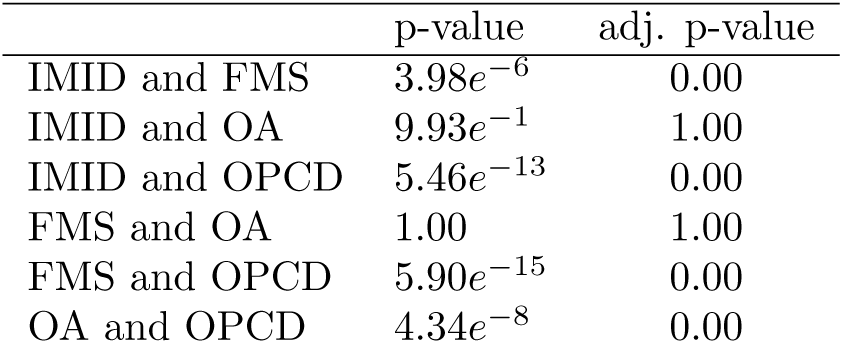
p-values and adjusted p-values obtained by Pearson’s *χ*^2^-test.

#### A.2.3 Results of MLC Models without OA

**Table A3:** Average performance of the MLC approaches with *C* = 3 classes, namely *FMS*, *OPCD* and *IMID*.

| MLC with $C = 3$<br>$\{FMS, OPCD, IMID\}$<br>no rebalancing | |
| --- | --- |
| DT |  |
| Sensitivity | 49.2 % |
| Specificity | 83.7 % |
| Precision | 65.7 % |
| F<sub>1</sub>-Score | 55.9 % |
| RF |  |
| Sensitivity | 51.9 % |
| Specificity | 86.1 % |
| Precision | 70.0 % |
| F<sub>1</sub>-Score | 59.2 % |
| LRM |  |
| Sensitivity | 49.4 % |
| Specificity | 86.4 % |
| Precision | 69.7 % |
| F<sub>1</sub>-Score | 57.1 % |
| MLP |  |
| Sensitivity | 54.1 % |
| Specificity | 78.9 % |
| Precision | 62.1 % |
| F<sub>1</sub>-Score | 57.1 % |
| $k$-NN | |
| Sensitivity | 41.4 % |
| Specificity | 86.3 % |
| Precision | 67.4 % |
| F<sub>1</sub>-Score | 50.5 % |

